# Multimodal imaging of microstructural cerebral changes and loss of synaptic density in Alzheimer’s disease

**DOI:** 10.1101/2023.04.14.23288516

**Authors:** Soodeh Moallemian, Eric Salmon, Mohamed Ali Bahri, Nikita Beliy, Emma Delhaye, Evelyne Balteau, Christophe Phillips, Christine Bastin

**Author notes:** Corresponding authors; Email addresses (Christophe Phillips), (Christine Bastin). Contributed equally (Christophe Phillips, Christine Bastin).

## Abstract

Multiple neuropathological changes are involved in Alzheimer’s disease (AD). The current study investigated the concurrence of neurodegeneration, increased iron content, atrophy, and demyelination in AD.

Quantitative multiparameter MRI maps providing neuroimaging biomarkers for myelination and iron content along with synaptic density measurements using [18F] UCB-H PET were acquired in 24 AD and 19 Healthy controls (19 males).

The whole brain voxel-wise group comparison revealed demyelination in the right hippocampus, while no significant iron content difference was detected. Bilateral atrophy and synaptic density loss was observed in the hippocampus and amygdala. The multivariate GLM (mGLM) analysis shows a bilateral difference in the hippocampus and amygdala, right pallidum, left fusiform and temporal lobe suggesting that these regions are the most affected despite the diverse changes in brain tissue properties in AD. Demyelination was identified as the most affecting factor in the observed differences.

Here, the mGLM is introduced as an alternative for multiple comparisons between different modalities, reducing the risk of false positives while informing about the co-occurrence of neuropathological processes in AD.

## 1 Introduction

Concomitantly with an increase in average life expectancy, a major epidemiologic trend of the current century is the rise of neurodegenerative diseases worldwide, among which Alzheimer’s disease (AD) is the most common type, with 60 to 80 percent of the cases (Calabrò et al., 2021). Despite an incipient decrease in incidence, AD prevalence is expected to rise because this neurodegenerative disease increases exponentially with age (Azam et al., 2021; C.-C Tan, 2014). It is recognized that AD pathological processes unfold decades before the emergence of clinical signs of cognitive decline (Dean et al., 2017; Gonneaud and Chételat, 2018; Tan et al., 2014). These pathological processes include a progressive accumulation of amyloid-beta plaques and tau neurofibrillary tangles (NFT), in addition to synaptic and neuronal loss (Gulisano et al., 2018; Jack et al., 2013; Spillantini and Goedert, 2013; Tan et al., 2014; Yin et al., 2021). According to the amyloid cascade hypothesis (Jack et al., 2013), amyloid plaques are the initial cause of AD, triggering tau NFT, synaptic and neuronal loss. Synaptic loss appears as the best correlate of cognitive decline in patients with Mild Cognitive Impairment (MCI) and with AD (DeKosky and Scheff, 1990; Scheff et al., 2006; Terry et al., 1991). Interestingly, in about 40% of cognitively normal older individuals, neurodegeneration (hippocampal atrophy) precedes detection of amyloid plaques (Jeremic et al., 2021; Villemagne et al., 2011). Accordingly, the identification of the early events in the AD pathophysiological cascade with in vivo noninvasive methods is critical to increase our understanding of the disease and inform the search for a treatment.

In the quest for early biomarkers, it has been suggested that changes in brain microstructure are among the first manifestations of AD (Bartzokis, 2011). Increased iron levels are associated with dysfunction of oligodendrocytes, notably impacting myelin repair. Moreover, an increase in free iron is toxic, inducing oxidative stress and inflammation, cell dysfunction, and, ultimately, cell death (Bartzokis, 2011; Bulk et al., 2018; Calabrò et al., 2021). So, it is hypothesized that myelin breakdown and increases in iron levels are very early events in the physiopathology of Alzheimer’s disease. In support of this “myelin and iron” hypothesis, histological studies showed that myelin breakdown in early AD occurs mainly in frontal and temporoparietal areas (Bartzokis, 2011; Bulk et al., 2018; Kalpouzos et al., 2017; Zecca et al., 2004). Increased iron content was also found in frontal and temporal areas of AD patients (Bulk et al., 2018; House et al., 2008). Moreover ex vivo studies showed that altered iron accumulation is positively correlated with the number of amyloid-beta plaques in these areas (Bulk et al., 2018; Duijn, 2017). Elevated iron content has also been observed in the hippocampus of AD patients (Zeineh et al., 2015). Additionally, higher levels of ferritin (i.e., the principal iron storage protein of the body) in the cerebrospinal fluid (CSF) are associated with the poorer cognitive performance of cognitively normal, MCI and AD participants, and predicted MCI conversion to AD (Ayton et al., 2015; Peng et al., 2021). Novel neuroimaging tools can be used to assess brain microstructure. Indeed, specific MRI parameters have differential sensitivity for structural aspects of tissue such as fiber coherence, macromolecules, myelin, iron, and water content. Recently developed quantitative MRI techniques offer, through their sensitivity to microstructural tissue properties, a unique opportunity for establishing in vivo the link to findings of postmortem histological assessment of brain tissue. Notably, Multi-Parameter Mapping (MPM) has been used to create quantitative brain maps that lead to a highly specific inference of tissue properties such as myelin water fraction (i.e., myelination) and iron content in the gray matter (Draganski et al., 2011). Consistently with ex vivo histological studies indicating degeneration of myelin sheaths with healthy aging (Peters, 2002), myelin water fraction as measured by MPM magnetization transfer saturation maps (MTsat) was found to decrease with aging in the corpus callosum as well as in frontal and parietal white matter (Callaghan et al., 2014). Additionally, in line with ex vivo evidence of increased iron content in basal ganglia in normal aging (Bulk et al., 2018), MPM imaging detected in vivo a positive correlation between iron deposit in the basal ganglia and age (Callaghan et al., 2014; Draganski et al., 2011). Increased iron content was related to lower blood oxygen level dependent (BOLD) signal in older adults (Kalpouzos et al., 2017). A more recent aging study on a large-scale cohort confirmed the age-related atrophy and demyelination, and reported an overlap between interregional volume and tissue property differences in aging that affected predominantly motor and executive networks (Taubert et al., 2020).

In this context, our main objective was to use quantitative MRI to detect in vivo microstructural changes (myelin water fraction and iron content) in individuals with AD characterized by significant amyloid burden in the brain by comparison to healthy older individuals (amyloid-negative and/or cognitively healthy). In AD, investigation of brain microstructure with quantitative MRI using MPM protocol has only recently started. One study (Acosta-Cabronero et al., 2016) used quantitative susceptibility mapping in MRI to show that AD patients have increased iron content in the putamen, caudate nucleus, and amygdala. The same authors (Acosta-Cabronero et al., 2013) indicated that in healthy older adults, iron accumulation can be found in frontal lobes, affecting brain regions related to motor functions. Steiger and colleagues observed a decrease in gray matter volume and myelin, and an increase of iron in widespread brain regions including the basal ganglia in older adults using quantitative MRI technique (Steiger et al., 2016). Another work directly evaluated, in healthy older participants, the concurrent relation between CSF markers of amyloid-beta and tau AD pathology, and MRI relaxometry-based measures of myelin content in the brain (Dean et al., 2017). They found that lower CSF amyloid-beta and higher tau levels were related to regional decreases in the brain MRI myelin measures, particularly in brain regions known to be preferentially affected in AD, including white matter in the frontal, temporal, corpus callosum, and cingulum regions.

To our knowledge, no study has used quantitative MPM to assess in vivo myelin and iron in AD concomitantly. Moreover, little is known about in vivo co-occurrence of cerebral microstructural changes and synaptic loss. The latter can be assessed with PET imaging using radiotracers binding to synaptic vesicle protein 2A (SV2A). With SV2A-PET imaging, AD-related decreased synaptic density was found in several cortical areas and the thalamus, with the most significant effect size in the hippocampus (Bastin et al., 2020; Chen et al., 2018).

In the current study, co-occurrence of microstructural changes with reduced regional cerebral uptake of [18F]UCB-H PET indexing synaptic density was assessed with a multivariate model applied to the different imaging modalities (MPM and SV2A-PET). If myelin decrease and iron burden are early events preceding synaptic loss and neuronal death (Bartzokis, 2011), one should observe a co-occurrence in microstructural changes and decreased synaptic density in the case of AD, likely in the hippocampus whose alteration drives symptoms in the patients (Bastin et al., 2020). Of note, in the current cross-sectional study, we cannot assess the chronology of pathological changes across the different modalities. Nevertheless, we hypothesize that, if pathological processes are triggering one another as suggested by Bartzokis (2011), one should observe co-localization of pathological changes in the brain.

## 2 Methods

### 2.1 Participants

The data come from a published study that focused on in vivo imaging of synaptic loss (Bastin et al., 2020). Two groups of older participants were included in the study. The first group consisted of 24 amyloid-positive patients from the AD continuum (Aβ-positive group), which encompasses individuals diagnosed with mild cognitive impairment (MCI) as well as those diagnosed with probable AD. These patients were recruited from the Memory Clinic at Liege University Hospital. They were diagnosed based on current NIA-AA criteria (Albert et al., 2011; Jack et al., 2018; McKhann et al., 2011). As part of the initial diagnostic process, [18F]FDG-PET was used as a biomarker of neurodegeneration in all patients. Also, global cognition was assessed with the Mini-mental state examination (MMSE). Aβ positivity was determined based on [18F]Flutemetamol-PET by qualitative visual inspection and by cortical standardized uptake value ratios (SUVR) above a quantitative threshold determined in a database of healthy older adults (Bastin et al., 2020). In Aβ-positive group, 5 patients were diagnosed with MCI (MMSE between 26 and 30) and 19 with probable AD, with MMSE scores between 14 and 26 (mild stage, MMSE > 20, n = 15; moderate stage, MMSE< 20, n = 4). The second group comprised 19 cognitively healthy controls (HC) (with MMSE between 28 and 30). In the HC group, amyloid-negativity was confirmed in eight participants. For participants who did not undergo an amyloid PET, because they refused an additional PET exam, they were considered healthy controls if they showed no or only minimal hippocampal atrophy on MRI, as assessed by visual inspection by a neurologist (Dubois et al., 2007). Both groups were matched for age, sex, and education. Table 1 shows a summary of the subjects.

**Table 1.**
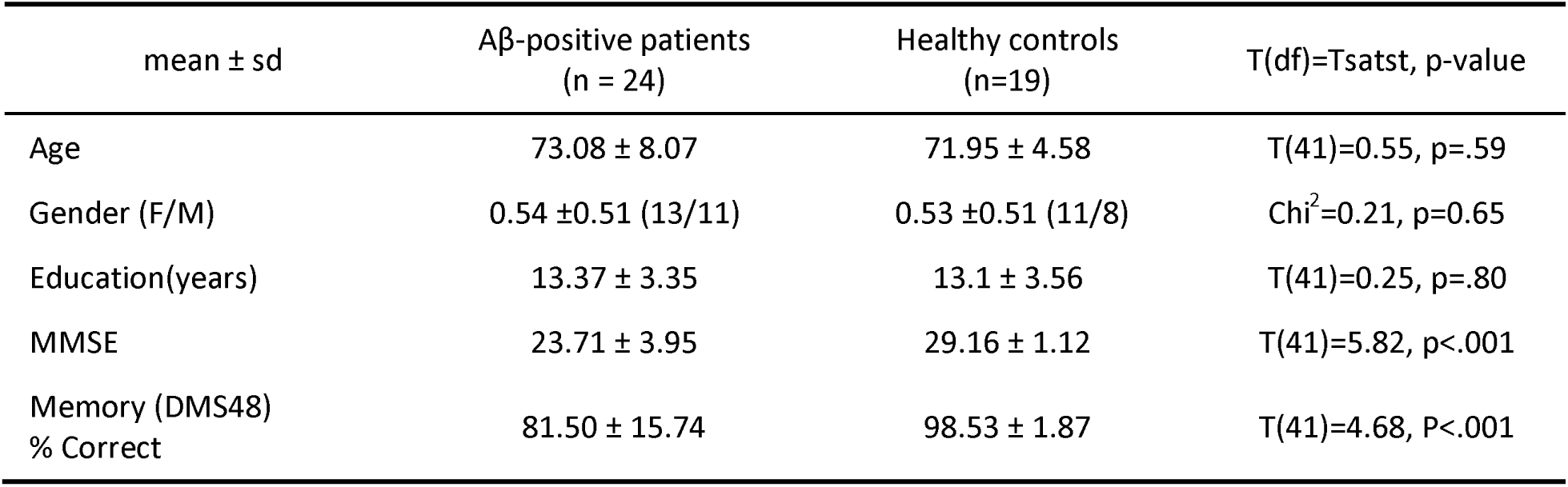
Demographics, clinical characteristics, and neuropsychological scores of AD and HC groups. Abbreviations: sd, standard deviation; Aβ, Amyloid beta; MMSE, mini mental state examination; DMS48, visual recognition memory, Tstat, T-student statistics, df, degrees of freedom, F, female, M, male.

| mean ± sd | Aβ-positive patients<br>(n = 24) | Healthy controls<br>(n=19) | T(df)=Tstat, p-value |
| --- | --- | --- | --- |
| Age | 73.08 ± 8.07 | 71.95 ± 4.58 | T(41)=0.55, p=.59 |
| Gender (F/M) | 0.54 ± 0.51 (13/11) | 0.53 ± 0.51 (11/8) | Chi<sup>2</sup>=0.21, p=0.65 |
| Education(years) | 13.37 ± 3.35 | 13.1 ± 3.56 | T(41)=0.25, p=.80 |
| MMSE | 23.71 ± 3.95 | 29.16 ± 1.12 | T(41)=5.82, p<.001 |
| Memory (DMS48)<br>% Correct | 81.50 ± 15.74 | 98.53 ± 1.87 | T(41)=4.68, P<.001 |

### 2.2 Data acquisition

Our data consists of dynamic PET and MRI

#### 2.2.1 SV2A-PET

Dynamic PET acquisitions were carried out using a Siemens/ CTI (Knoxville, TN) ECAT HR+ PET scanner. An intravenous bolus of [18F]UCB-H[37] of 157.06 ± 8.96 MBq was administered. For a total of 100 minutes, the dynamic PET was conducted with time frames of 6*10s, 8*30s, 5*120s, and 17*300s. All PET images were reconstructed using filtered back projection (Hann filter, 4.9 mm FWHM), including corrections for measured attenuation, dead time, random events, and scatter using standard software (ECAT 7.1, Siemens/CTI, Knoxville, TN). The transaxial resolution in water, under these acquisition and reconstruction conditions, is 6.5–7 mm (voxel size 2.57 x 2.57 x 2.43 mm^3^). A mean unchanged plasma fraction was calculated for each group and used for modeling based on blood samples collected in 7 controls and 6 patients. Further information on PET acquisition and processing can be found in (Bastin et al., 2020).

#### Multi-parametric mapping MRI

MRI data has been acquired on a 3T whole-body MRI-scanner (Magnetom Prisma, Siemens Medical Solution, Erlangen, Germany) using a standard 32-channel head receiving coil. The whole-brain MRI acquisitions included a multiparameter mapping protocol (MPM) (Weiskopf et al., 2013). This protocol allows the estimation of (semi)quantitative maps for various parameters, including magnetization transfer saturation (MTsat), proportional to myelin; proton density (PD), proportional to water content; transverse relaxation (R2*), proportional to iron; and effective longitudinal relaxation (R1). The MPM protocol consists of 3 co-localized 3D multi-echo fast low angle shot (FLASH) acquisitions with 1 mm isotropic resolution and 2 additional calibration sequences to correct for inhomogeneities in the RF transmit field (Lutti et al., 2010). The FLASH datasets were acquired with predominantly PD, T1 and MT weighting determined by the repetition time (TR = 24.5 ms) and flip angle (FA = 6° for PD & MT, 21° for T1), referred to in the following as PDw, T1w and MTw echoes. MTw contrast was obtained using an additional off-resonance Gaussian-shaped RF pulse with 4 ms duration and 220 nominal flip angle, 2 kHz off-resonance before nonselective excitation. A high readout bandwidth of 320 Hz/pixel was used to minimize off-resonance and chemical shift artifacts (Helms and Dechent, 2009). Volumes were acquired in 176 sagittal slices using a 256×224 voxel matrix. GRAPPA parallel imaging was combined with partial Fourier acquisition to speed up acquisition time to approximately 20 min. Gradient echoes were acquired with alternating readout gradient polarity at 6 equidistant echo times [2.34, 4.68, 7.02, 9.36, 11.7, 14.04] ms. Two additional echoes were acquired for the PDw and T1w acquisitions at 16.38 ms and 18.72 ms.

B1 field mapping images (transmit B1+ and receive B1-fields) were also acquired to reduce spatial heterogeneities related to B1 effect, which was essential for proper quantification of T1 (or R1=1/T1) in particular. Finally, B0 field mapping images were acquired for image distortions correction: two magnitude images acquired at 2 different TE’s, and pre-subtracted phase images.

### 2.3 Image data processing

Collected data were anonymized and organized according to the Brain Imaging Data Structure (BIDS) (Gorgolewski et al., 2016) and its extensions for PET (Knudsen et al., 2020) and qMRI (Karakuzu et al., 2022) data, the latter using BIDSme (https://github.com/CyclotronResearchCentre/bidsme), and the former with custom MATLAB scripts. All information needed for subsequent analysis was incorporated into the dataset. Data is available from the corresponding authors upon reasonable request.

#### 2.3.1 MRI

To obtain the quantitative maps, MRI data were processed with SPM12 (www.fil.ion.ucl.ac.uk/spm) and the hMRI (https://hmri.info/) toolbox, where the latter is an extension to SPM (Tabelow et al., 2019). T1w, PDw, and MTw images acquired at multiple TEs were extrapolated to TE=0 to increase the signal-to-noise ratio and remove the otherwise remaining R2* bias (Tabelow, 2019). The TE=0 extrapolated MTw, PDw, and T1w images were used to calculate MT saturation, R1 and apparent signal amplitude A* maps. A* maps were rescaled to generate PD maps. All quantitative maps were corrected for inhomogeneities from local RF transmit field (B1+), using B1 and B0 field mapping images (Lutti et al., 2010). The receive bias field map (B1-) was used to correct PD maps for instrumental biases (Ashburner and Friston, 2005). R2* maps were estimated using the ESTATICS method from the three different FLASH acquisitions by accounting for the varying contrasts. The ordinary least squares (OLS) log-linear fit was also used to detect and down weight echoes affected by motion artifacts (Weiskopf et al., 2014). R1 maps were corrected for the radio frequency (RF) transmit field inhomogeneity B1+ (Preibisch and Deichmann, 2009). Quantitative maps were segmented into gray matter (GM), white matter (WM), and cerebrospinal fluid (CSF) using the unified segmentation approach as implemented in SPM (Ashburner and Friston, 2005). Inter-subject registration of the GM and WM tissue maps was performed using DARTEL, a nonlinear diffeomorphic algorithm (Ashburner, 2007). This algorithm estimates the deformations that best align the tissue probability maps by iterative registration of these maps to their average. The tissue probability maps were then normalized to the stereotactic space specified by the Montreal Neurological Institute (MNI) template using the resultant DARTEL template and deformations. Then, for voxel-based morphometry (VBM) analysis, specific tissue-weighted smoothing, with a 3mm FWHM isotropic kernel, was applied to avoid mixing values from different tissues classes, as would happen with standard Gaussian smoothing.

A GM mask was created using the mean segmented MTsat image from all subjects to be later used as an explicit mask in the statistical analysis.

#### 2.3.2 PET

PET data were processed as described previously (Bastin et al., 2020). In brief, [18F]UCB-H PET dynamic frames were corrected for motion without re-slicing. The images were corrected for partial volume effects (PVE) using the iterative Yang voxel-wise method implemented in the PETPVC toolbox (Thomas et al., 2016), with GM, WM, CSF and “other” as ROI masks. Kinetic modeling using PVE-corrected dynamic PET data and image-derived input function was done with PMOD (Version 3.7, PMOD Technologies, Zurich, Switzerland). Input function was derived from the dynamic images (Bahri et al., 2017) and corrected for metabolites using the measured group mean unchanged plasma fraction. Logan graphical analysis (with t* = 25 min) was used to calculate the distribution volume (Vt) map of [18F]UCB-H in the brain. Finally individual Vt maps were coregistered with their corresponding MTsat map, then their spatial normalization transformations were applied to warp the Vt maps in the same reference space.

### 2.4 Statistical analyses

All analyses focused on GM only as PET images indexing synaptic density are only interpretable for gray matter. Therefore, an explicit mask for GM was applied on all the analysis. Since each parametric map has a specific unit, e.g., Hertz for R2* images and ml/cc for Vt maps, their intensities are not directly comparable. Thus, all maps were Z-transformed - per modality and across subjects - using the grand mean and variance over each voxel, to ensure comparability of different modalities for our multivariate analysis. All statistical analyses were performed on standardized data.

For quantitative MRI, we decided not to investigate R1 and PD maps, as they are proportional to multiple tissue properties at the same time, and would lead to underestimation of microstructural changes that we are interested in.

Three quantitative modalities (MTsat, R2*, and Vt) and GM density maps were individually analyzed using a univariate 2-sample t-test GLM with age and sex of the participants as covariates. We tested the difference between the two groups for each modality. The t-student contrast defined for MTsat, Vt, and GM volume (GMvol) maps was HC>AD, hypothesizing that healthy controls have more myelin, synapses, and GM volume than AD patients. For R2* maps, we used contrasted AD>HC, hypothesizing that R2* values in AD group are superior to those of healthy subjects, as increased iron load is considered toxic. A MANOVA model was specified using the design matrixes of the three univariate models in the MSPM toolbox (Gyger et al., 2021), a newly developed toolbox working under SPM as a multivariate extension of univariate GLM (Chen et al., 2014; M Mcfarquhar, 2016). The multivariate GLM (mGLM) models the multivariate observations as Y=XB+E, where Y_43×4_ =[Y_1_,Y_2_,Y_3_,Y_4_J is the multi-modal data matrix, each row of Y represents one subject, and each column of Y represents one modality MTsat, R2*, GMvol, and Vt at a single voxel; and X_43×4_ = [X_1_,X_2_,X_3_,X_4_Jis the design matrix, representing the AD and HC groups in the first two columns, and age and sex of the participants in the last two columns. The matrix B is a 4 × 4 matrix of size of regression coefficients; and E is the residual matrix of size 43 × 4. Matrix B is estimated using an ordinary least square method.

The regression model can be used to partition the total variation in the outcome into explained variance and unexplained variance. In this sense, the total sums of square and cross products (SSCP) terms are calculated as: SSCP_Total_=SSCP_Model_+SSCP_Residual_. The SSCP matrix is used to estimate the variance-covariance matrix of the predictor variables in linear regression analysis, and can be presented as 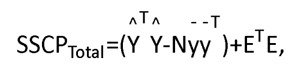 where in our case N=4, the total number of observations. To test the null hypothesis that all the coefficients in B are equal to zero, we can compute the eigenvalues of SSCP_Model_SSCP_Residual_. Then the Wilk’s lambda summary statistics Λ is calculated based on the eigenvalues solving the equation of eigen-decomposition for the determinant matrix.

Here, we determine the canonical vectors for our test statistic. In this context, canonical vectors refer to the linear combinations of the original variables that maximize the separation between groups or conditions in the multivariate space (Tabachnick and Fidell, 2007). These vectors are determined by performing a multivariate analysis of variance (MANOVA) and extracting the canonical variates. The F-test is then applied to test the significance of the overall multivariate effect. Canonical vectors provide insights into the relationships between variables and facilitate the identification of the most influential factors driving group differences. For illustrative purposes only, we extracted the original values, after Z-transformation, from the MTsat, R2*, GMvol, and Vt maps used at the voxels within significant clusters, which corresponds to the difference in real tissue property values.

To test the hypothesis of the association between all dependent variables and contrasts among predictors, we applied this linear hypothesis: H_0_: CBL = 0, where L_4×4_ is a full rank matrix to test the hypothesis, here an identity matrix to test the hypothesis for the joint effect of all modalities (columns of B). The contrast matrix C=l1,-1,0,0,0J would perform a standard F-test to assess the difference between the 2 groups, AD and HC. See (Tabachnick and Fidell, 2007) for mathematical details. Additional group comparison was performed on MTsat, R2*, GMvol, and Vt maps after adjustment for age and gender covariates and masking for the significant ROIs resulted from the mGLM model. Correlation analyses between memory, age, and sex with different modalities, was performed on the AD and HC group.

## 3 Results

Statistical inference was performed using a p-value < .05 “family-wise error rate” (FWER) corrected, at the voxel or cluster extent levels. For the latter, the cluster forming threshold used was a voxel level p<.001 uncorrected.

### 3.1 Univariate analyses

Coordinates and anatomical labels of the peaks are presented in Table 2, showing the voxel-wise comparisons of two groups of AD and HC for different univariate analysis performed on MTsat, R2*, GMvol, and Vt maps.

**Table 2.** Significant differences between AD participants and healthy controls for MTsat and Vt maps. Brain regions were labeled with the AAL3 atlas toolbox in SPM. This table shows up to 3 peaks (at least 8mm apart) within each cluster. Clusters were thresholded to contain >20 voxels. Coordinates are MNI coordinates. FWER correction was applied for P<0.05 at cluster level. Clusters highlighted with *, were significant at voxel level (P<.05). Abbreviations: FWER, family-wise error rate, MTsat, magnetization transfer saturation, Vt, total volume distribution, GM, gray matter, df, degree of freedom.

| peak [x y z]<br>Coordinates | Cluster<br>P-value | Cluster Size<br>#voxels | Brain Region |
| --- | --- | --- | --- |
| MTsat (df=39) |  |  |  |
| [31 -7 -27] | 0.001 | 2508 | Right Para Hippocampal |
| [37 -21 -12] |  |  | Right Hippocampus |
| [20 -11 -20] |  |  | Right Amygdala |
| GM volume (df=39) |  |  |  |
| [30 1 -34] | 0.000 | 5855 | Right Hippocampus |
| [21 -1 -16] |  |  | Right Amygdala |
| [31 -24 -15] |  |  | Right Para Hippocampal |
| [-21 -1 -16] | 0.000 | 4848 | Left Amygdala |
| [-27 -11 -16] |  |  | Left Hippocampus |
| [-25 -4 -38] |  |  | Left Fusiform |
| [-24 -23 -23] | 0.000 | 966 | Left Hippocampus |
| [-40 -51 -15] | 0.020* | 30 | Left fusiform |
| [6 5 -12] | 0.009* | 27 | Left Olfactory blub |
| [-38 -12 12] | 0.012* | 57 | Left Insula |
| [5 34 31] | 0.019* | 34 | Right Mid. Cingulate |
| [-7 38 14] | 0.016* | 41 | Left Pre. ACC |
| [52 -46 -16] | 0.015* | 47 | Right Inf. Temporal |
| [36 -33 -23] | 0.01* | 69 | Right Fusiform |
| Vt (df=39) |  |  |  |
| [25 -11 -15] | 0.000* | 1462 | Right Hippocampus |
| [30 -9 -21] |  |  | Right Amygdala |
| [17 -7 -15] |  |  | Right Para Hippocampal |
| [40 49 22] | 0.008* | 137 | Right Mid. Frontal |
| [-27 -12 -16] | 0.000* | 1506 | Left Hippocampus |
| [-22 -36 4] |  |  | Left Thalamus |
| [3 -6 8] | 0.000 | 200 | Right Thalamus |

#### 3.1.1 Magnetization transfer saturation (MTsat)

MTsat maps revealed a significant difference at cluster-level between AD patients and healthy controls after correcting for FWER (P < .05) which covers the right hippocampus and amygdala indicating lower values in AD than in controls. Figure 2-A shows the statistical parametric map of univariate analysis of MTsat maps at p <.001 uncorrected.

**Figure 1.**
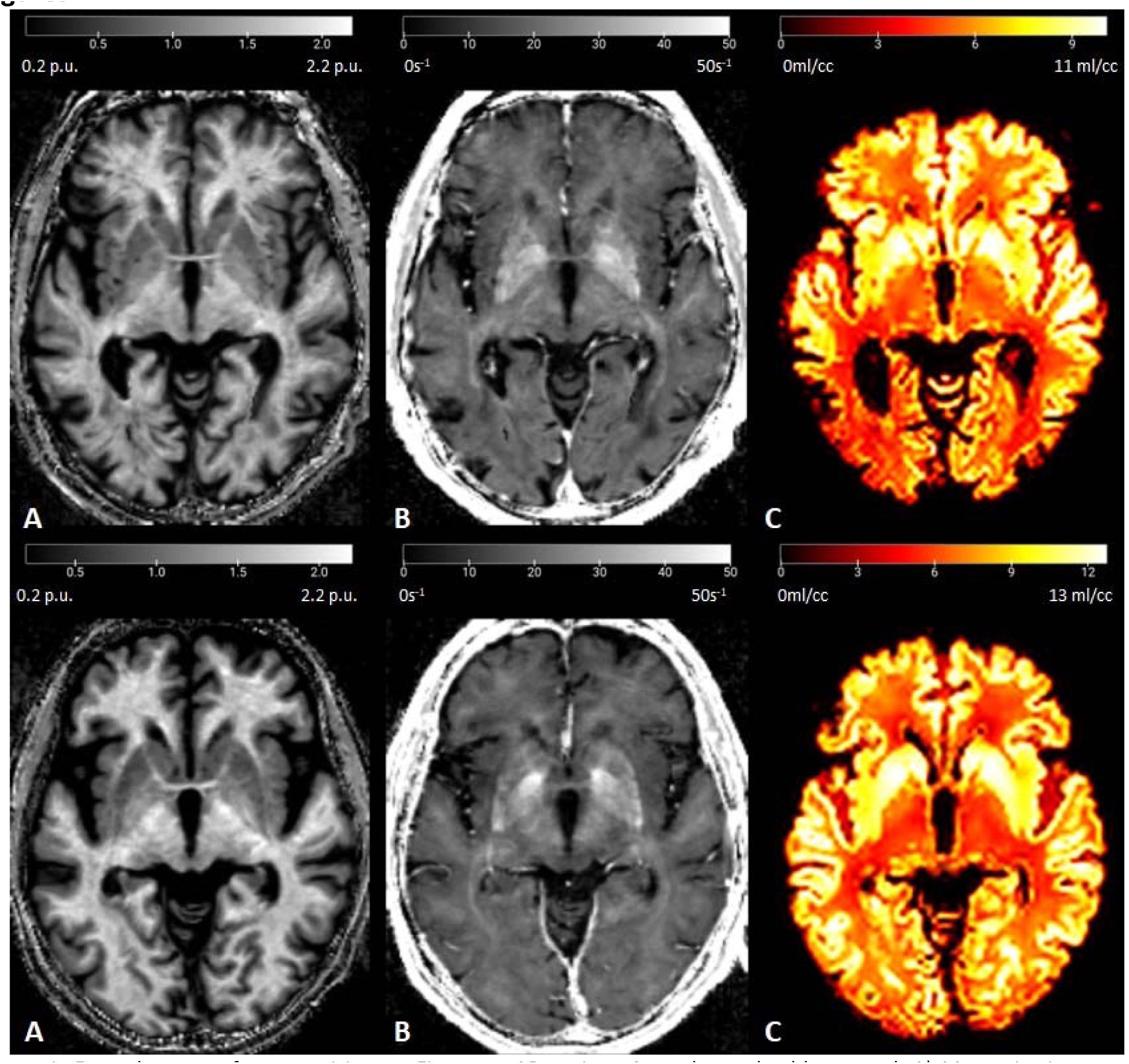
Example maps of two participants. First row, AD patient, Second row, healthy control. A) Magnetization transfer saturation, MTsat; B) Effective transverse relaxation rate, R2*; C) Total volume distribution, Vt.

**Figure 2.**
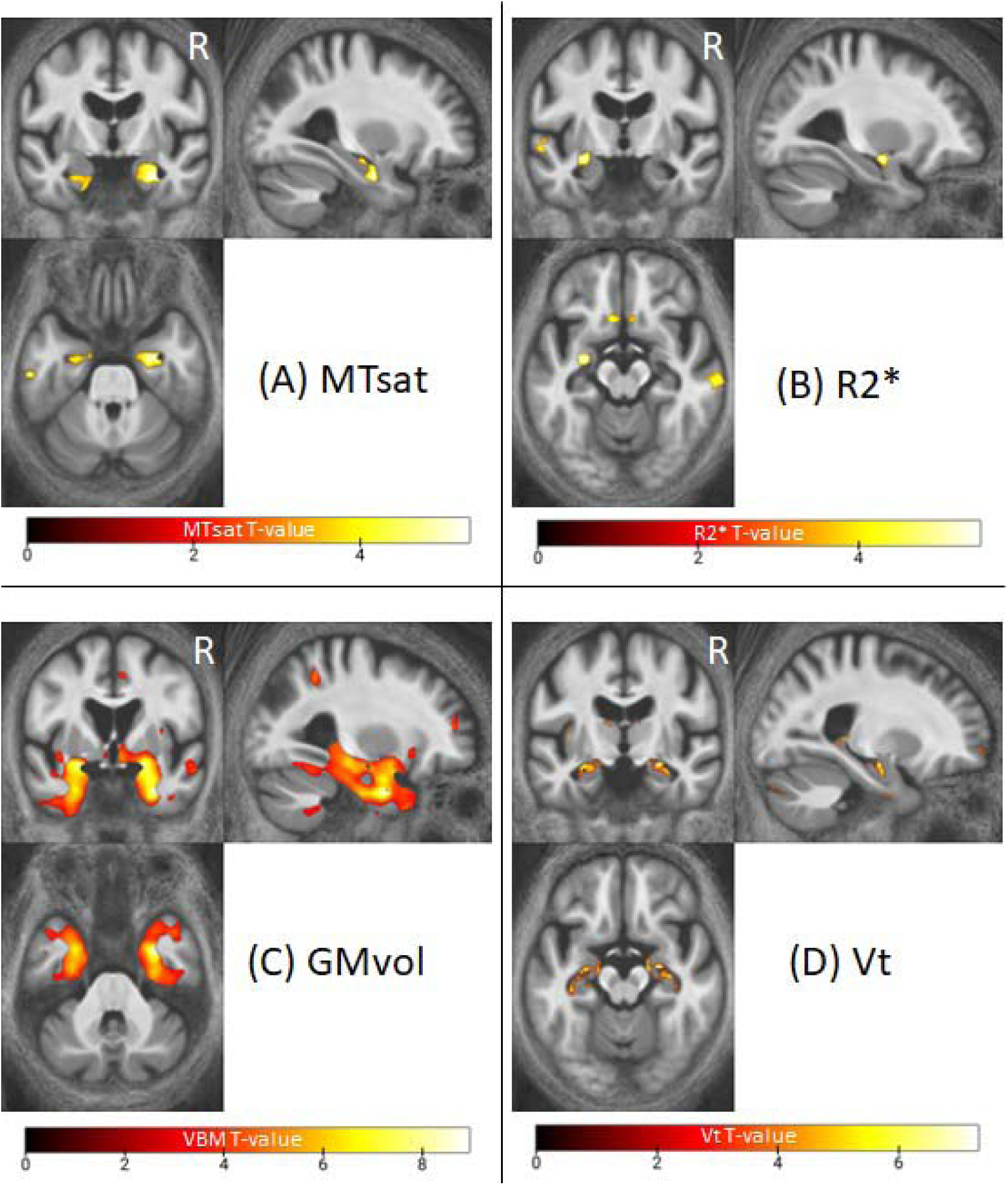
Statistical parametric maps of the univariate analysis for the difference between AD and HC groups. The SPMs were super imposed on the mean MTsat map for both groups on MNI space. For illustration purposes displayed at p<.001 uncorrected for FWER. Abbreviations: MTsat, magnetization transfer saturation; R2*, effective transverse relaxation rate; gray matter volume; Vt, total volume distribution; FWER, family-wise error rate.

#### 3.1.2 Effective transverse relaxation rate (R2*)

No significant difference in R2* maps, representative of iron level content in the brain, was detected between the groups. However, at a more lenient statistical threshold (p < .001 uncorrected), the results for R2* analysis show differences in the superior part of orbitofrontal cortex bilaterally as well as in the left hippocampus and right mid-temporal gyrus (see Figure 2-B).

#### 3.1.3. Voxel-based morphometry

We could identify significant reduction in GM volume in AD group (p <.05, corrected for FWER) bilaterally in hippocampus and fusiform cortex, and in left amygdala, olfactory bulb, and anterior cingulate cortex (Figure 2-C).

#### 3.1.4 Total [18F]UCB-H volume distribution

Vt data shows higher intensities in healthy controls compared to AD patients in the left and right hippocampus and amygdala at voxel-level (P_FWE_ <.05) as well as, right and left thalamus (Figure 2-D).

#### 3.1.5 Correlation analyses

The results from Pearson’s correlation analyses in the AD group are presented in Table 5. There were significant correlations between MTsat and GMvol maps (r = .462, p<.05) in right parahippocampal cortex. A strong correlation was also observed in the left fusiform between MTsat and Vt maps (r = .724, p<.001). R2* and GMvol maps were negatively correlated in right parahippocampal cortex (r = -.574, p<.01) and left fusiform (r = -.628, p<.01). GMvol and Vt maps were found positively correlated in left hippocampus (r = .472, p< .05) and fusiform (r = .447, p<.05).

In the AD group, there were no significant correlations between memory (indexed by DSM48 score) and the different imaging modalities. Moreover, MTsat showed a negative correlation with age and gender in left and right hippocampus of AD participants (r =-.431, r = -.445, p< .05), as well as in left fusiform and temporal clusters.

Detailed correlation results for age and gender are presented in the supplementary data (Table sup-1 and sup-2).

### 3.2 Multivariate analysis

The multivariate GLM (mGLM) model, controlling for the effect of age and sex, shows significant difference at voxel-level (P_FWE_ <.05) between the two groups in both left and right hippocampus and amygdala, left fusiform and superior temporal gyrus, and right para hippocampal (Figure 3). Coordinates and anatomical labels of the peaks are presented in Table 3. We also investigated the multicollinearity between different maps in the significant clusters of the mGLM model. The variance inflation factor (VFI) in all cases is in the [0.98, 1.3] range, suggesting no multicollinearity.

**Figure 3.**
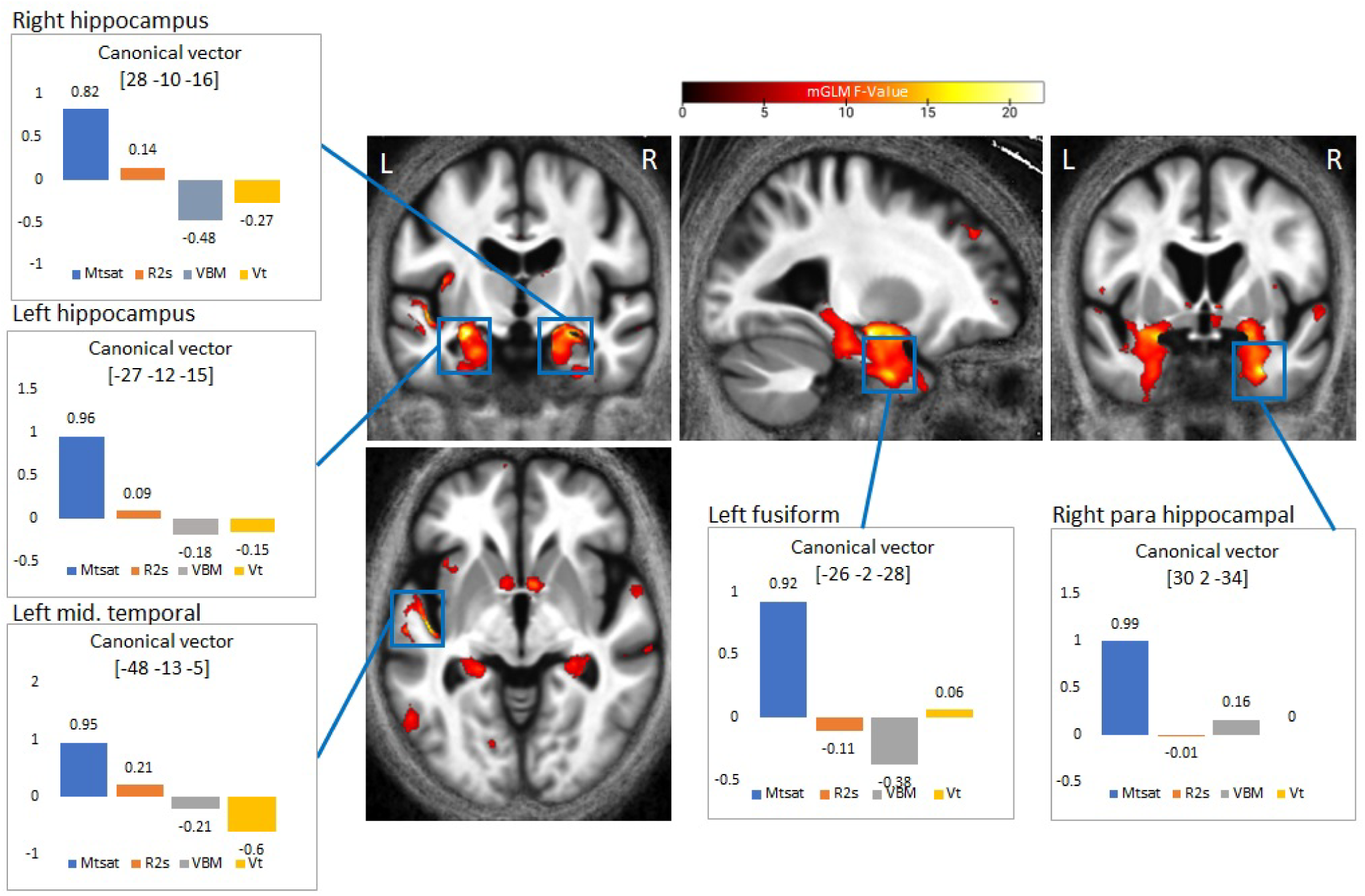
Statistical parametric map of the multivariate analysis for the difference between AD and HC groups. The mSPM was super imposed on the mean MTsat map for both groups in MNI space. For illustration purposes displayed at p<.001 uncorrected for FWER. Canonical vectors for the peak voxel of each cluster in Table3 are depicted in colored bars (blue=MTsat, orange=R2*, gray, GM vol., yellow=Vt) with arbitrary units. Abbreviations: MTsat, magnetization transfer saturation; R2s, effective transverse relaxation rate; GMvol, gray matter volume; Vt, total volume distribution; FWER, family-wise error rate.

**Table 3.** Significant differences between AD and HC groups in the mGLM model. Brain regions were labeled with the aal3 atlas toolbox in SPM. This table shows up to 3 peaks (at least 8mm apart) within each cluster. Clusters were thresholded to contain >20 voxels. FWER correction was applied for P<.05 at cluster level. Clusters highlighted with *, were significant at voxel level (P<0.05). Abbreviations: FWER, family-wise error rate, df, degree of freedom, mGLM, multivariate GLM, AD, Alzheimer’s disease, HC, healthy controls.

| MSPM (df=36) |  |  |  |
| --- | --- | --- | --- |
| peak [x y z]<br>Coordinates | Cluster<br>P-value | Cluster Size<br>#voxels | Brain Region |
| [-27 -12 -15] | 0.000* | 569 | Left Hippocampus |
| [-22 0 -16] |  |  | Left Amygdala |
| [28 -10 -16] | 0.000* | 707 | Right Hippocampus |
| [19 -2 -16] |  |  | Right Pallidum |
| [22 -8 -21] |  |  | Right Amygdala |
| [30 2 -34] | 0.000* | 111 | Right Para Hippocampal |
| [-26 -2 -38] | 0.002* | 54 | Left Fusiform |
| [-48 -13 -5] | 0.006* | 30 | Left Sup. Temporal |

#### 3.2.1 Canonical vectors

Canonical vectors are evaluated for the mGLM model for each modality. The canonical vectors at the peak voxel of significant clusters presented in Table 3 are illustrated in Figure 3 with colored bars. They suggest that MTsat maps were the most contributing factor to the observed difference between AD and HC groups.

The violin plots in Figure 4 represent the distribution of mean voxel values across subjects within different modalities and the significant clusters from the mGLM model in the original maps (after Z-transformation and correcting for age and gender). Group comparison for each ROI was computed with Student t test (p <0.05) as presented in Table 4. A significant difference was detected in the left fusiform and temporal for MTsat maps. In R2* maps, we could indicate a statistically significant difference in the left hippocampus. GM volume was significantly different in all the selected ROIs between the groups. Significant differences were observed in the right para hippocampal and left temporal.

**Figure 4.**
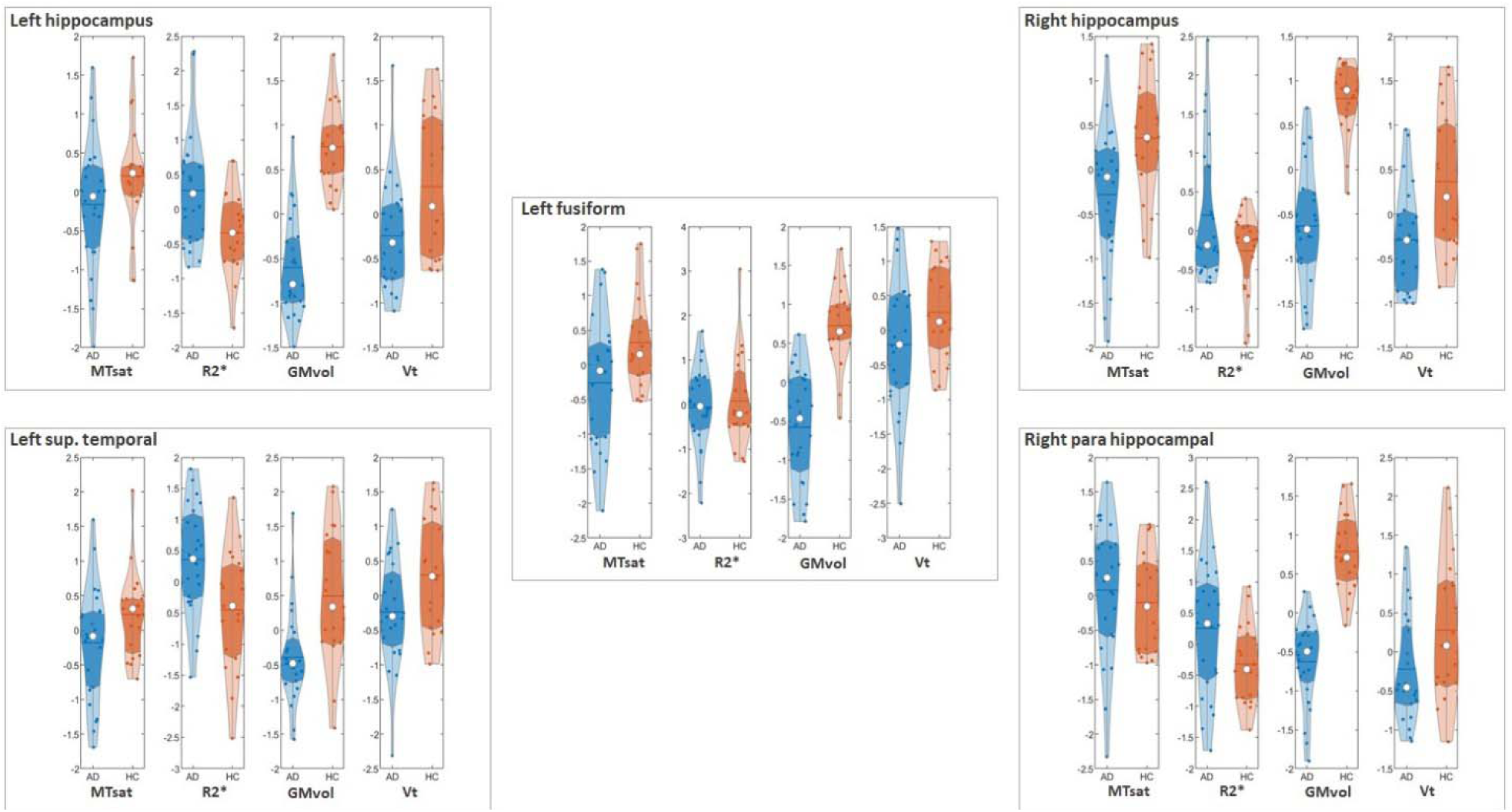
Raw voxel distribution of Z-transformed maps within each mGLM significant cluster. Each violin plot represents the distribution of the mean values across subjects within each AD (in blue) and HC (in red) cohort in the original map, after adjusting for age and gender. White circles represent the median value within each group per cluster. The range of y axes is based on the variations within each modality. The first and third quartiles are shown with darker shadows within each violin. The mean value within each group per modality and cluster is shown with the horizontal line. Abbreviations: MTsat, magnetization transfer saturation; R2s, effective transverse relaxation rate; GMvol, gray matter volume; Vt, total volume distribution; mGLM, multivariate GLM.

**Table 4.** ROI based voxel-level group comparison in MTsat, R2*, GM volume, Vt maps. The ROIs were selected from the statistical parametric map of the mGLM model. All maps were masked for these ROIs, then T-tests were performed for confidence interval of .95% on the adjusted values for age and gender. Abbreviations: MTsat, magnetization transfer saturation; R2*, effective transverse relaxation rate; Vt, total volume distribution; GMvol, gray matter volume; mGLM, multivariate GLM; df, degrees of freedom; diff, difference; sd, standard deviation.

| Modality | T(df)=tstat | P_value | Mean_diff ± sd | Cohen's d |
| --- | --- | --- | --- | --- |
| **Left hippocampus, #voxels per cluster = 569** |  |  |  |  |
| MTsat | T(41)= -1.6 | P = .12 | -.44 ± 0.90 | .49 |
| R2* | T(41)= 2.53 | P = .01 | .59 ± 0.74 | .79 |
| GMvol | T(41)= -6.93 | P < .001 | -1.38 ± 0.65 | 2.14 |
| Vt | T(41)= -2.68 | P = .01 | -.58 ± 0.71 | .81 |
| **Right hippocampus, #voxels per cluster = 707** |  |  |  |  |
| MTsat | T(41)= -2.71 | P < .001 | -.70 ± .82 | .83 |
| R2* | T(41)= 1.87 | P = 0.07 | .44 ± .75 | .59 |
| GMvol | T(41)= -7.50 | P < .001 | -1.47 ± .62 | 2.34 |
| Vt | T(41)= -3.19 | P < .001 | -.69 ± .71 | .96 |
| **Right para hippocampal, #voxels per cluster = 111** |  |  |  |  |
| MTsat | T(41)= .50 | P = .62 | .14 ± .91 | .16 |
| R2* | T(41)= 2.15 | P = 0.38 | .59 ± .87 | .68 |
| GMvol | T(41)= -7.16 | P < .001 | -1.47 ± .67 | 2.20 |
| Vt | T(41)= -2.09 | P = .04 | -.51 ± .81 | .63 |
| **Left fusiform, #voxels per cluster = 54** |  |  |  |  |
| MTsat | T(41)= -2.17 | P = .04 | -.62 ± .90 | .68 |
| R2* | T(41)= -.44 | P = .66 | -.13 ± .99 | .13 |
| GMvol | T(41)= -5.97 | P < .001 | -1.35 ± .72 | 1.86 |
| Vt | T(41)= -1.80 | P = .07 | -.49 ± .87 | .57 |
| **Left Sup. Temporal, #voxels per cluster = 30** |  |  |  |  |
| MTsat | T(41)= -1.72 | P = .09 | -.51 ± .95 | .54 |
| R2* | T(41)= 2.78 | P = .09 | .79 ± .93 | .85 |
| GMvol | T(41)= -3.46 | P < .001 | -.95 ± .91 | 1.04 |
| Vt | T(41)= -2.27 | P = .03 | -.59 ± .85 | .69 |

**Table 5.**
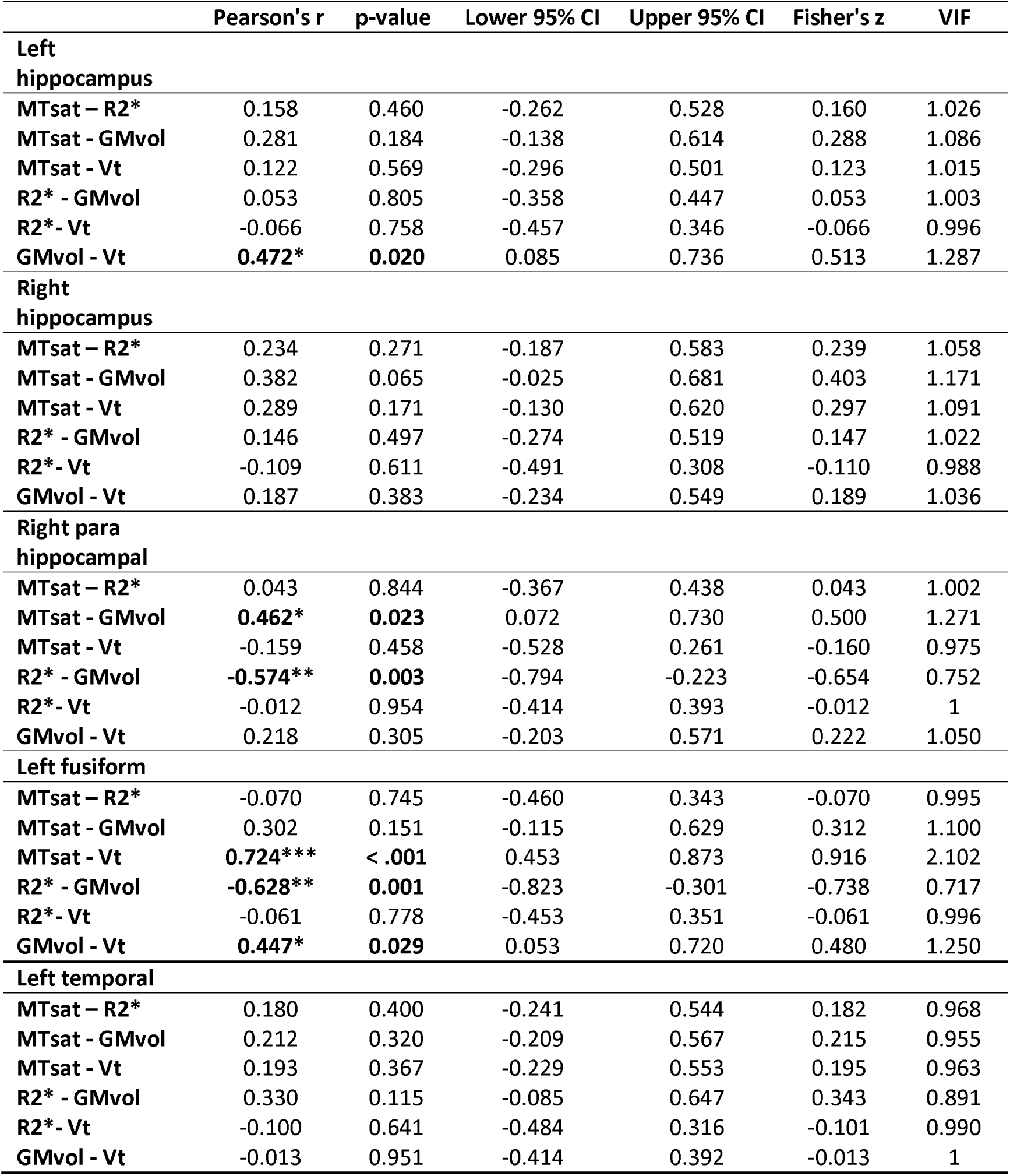
Pearson’s correlations between different maps in AD group. Significant correlations are highlighted by * p < .05, ** p < .01, *** p < .001. Abbreviations: CI, confidence interval; MTsat, magnetization transfer saturation; Vt, total volume distribution; R2*, effective transverse relaxation rate, GMvol, gray matter volume.

|  | Pearson's r | p-value | Lower 95% CI | Upper 95% CI | Fisher's z | VIF |
| --- | --- | --- | --- | --- | --- | --- |
| **Left hippocampus** |  |  |  |  |  |  |
| MTsat – R2* | 0.158 | 0.460 | -0.262 | 0.528 | 0.160 | 1.026 |
| MTsat - GMvol | 0.281 | 0.184 | -0.138 | 0.614 | 0.288 | 1.086 |
| MTsat - Vt | 0.122 | 0.569 | -0.296 | 0.501 | 0.123 | 1.015 |
| R2* - GMvol | 0.053 | 0.805 | -0.358 | 0.447 | 0.053 | 1.003 |
| R2* - Vt | -0.066 | 0.758 | -0.457 | 0.346 | -0.066 | 0.996 |
| GMvol - Vt | **0.472\*** | **0.020** | 0.085 | 0.736 | 0.513 | 1.287 |
| **Right hippocampus** |  |  |  |  |  |  |
| MTsat – R2* | 0.234 | 0.271 | -0.187 | 0.583 | 0.239 | 1.058 |
| MTsat - GMvol | 0.382 | 0.065 | -0.025 | 0.681 | 0.403 | 1.171 |
| MTsat - Vt | 0.289 | 0.171 | -0.130 | 0.620 | 0.297 | 1.091 |
| R2* - GMvol | 0.146 | 0.497 | -0.274 | 0.519 | 0.147 | 1.022 |
| R2* - Vt | -0.109 | 0.611 | -0.491 | 0.308 | -0.110 | 0.988 |
| GMvol - Vt | 0.187 | 0.383 | -0.234 | 0.549 | 0.189 | 1.036 |
| **Right para hippocampal** |  |  |  |  |  |  |
| MTsat – R2* | 0.043 | 0.844 | -0.367 | 0.438 | 0.043 | 1.002 |
| MTsat - GMvol | **0.462\*** | **0.023** | 0.072 | 0.730 | 0.500 | 1.271 |
| MTsat - Vt | -0.159 | 0.458 | -0.528 | 0.261 | -0.160 | 0.975 |
| R2* - GMvol | **-0.574\*\*** | **0.003** | -0.794 | -0.223 | -0.654 | 0.752 |
| R2* - Vt | -0.012 | 0.954 | -0.414 | 0.393 | -0.012 | 1 |
| GMvol - Vt | 0.218 | 0.305 | -0.203 | 0.571 | 0.222 | 1.050 |
| **Left fusiform** |  |  |  |  |  |  |
| MTsat – R2* | -0.070 | 0.745 | -0.460 | 0.343 | -0.070 | 0.995 |
| MTsat - GMvol | 0.302 | 0.151 | -0.115 | 0.629 | 0.312 | 1.100 |
| MTsat - Vt | **0.724\*\*\*** | **< .001** | 0.453 | 0.873 | 0.916 | 2.102 |
| R2* - GMvol | **-0.628\*\*** | **0.001** | -0.823 | -0.301 | -0.738 | 0.717 |
| R2* - Vt | -0.061 | 0.778 | -0.453 | 0.351 | -0.061 | 0.996 |
| GMvol - Vt | **0.447\*** | **0.029** | 0.053 | 0.720 | 0.480 | 1.250 |
| **Left temporal** |  |  |  |  |  |  |
| MTsat – R2* | 0.180 | 0.400 | -0.241 | 0.544 | 0.182 | 0.968 |
| MTsat - GMvol | 0.212 | 0.320 | -0.209 | 0.567 | 0.215 | 0.955 |
| MTsat - Vt | 0.193 | 0.367 | -0.229 | 0.553 | 0.195 | 0.963 |
| R2* - GMvol | 0.330 | 0.115 | -0.085 | 0.647 | 0.343 | 0.891 |
| R2* - Vt | -0.100 | 0.641 | -0.484 | 0.316 | -0.101 | 0.990 |
| GMvol - Vt | -0.013 | 0.951 | -0.414 | 0.392 | -0.013 | 1 |

The multivariate approach results for the difference in the right hippocampus go along with those of the univariate analyses of MTsat, GMvol, and Vt maps; narrowing the regions in which different micro-and macrostructural changes coincide.

## 4 Discussion

In this study, we investigated the association of myelination, iron accumulation, gray matter volume, and synaptic density, in patients with Alzheimer’s disease (i.e., presence of cerebral amyloid burden) compared to healthy individuals (with no/minimal hippocampal atrophy) in univariate and multivariate manner to discuss the co-occurrence of these changes at voxel level.

The three different microstructure markers are assumed to play a role during AD, with a hypothesized cascade of pathological processes whereby increased iron and decreased myelin would lead to neuronal death and a reduction of synapses (Bartzokis, 2011). Only a longitudinal study could capture this hypothetical chronology. Nevertheless, we hypothesized that, when reaching the clinical stage where amyloid-positive individuals demonstrate symptoms of cognitive decline, co-occurrence of changes in the myelination, iron level, and synaptic density should be seen if they were linked in previous stages. On the other perspective, macrostructural tissue loss has proved to be a sensitive marker for neurodegeneration despite its poor pathologic specificity (Mc Donald et al., 2010). Macrostructural atrophy was observed where microstructural abnormalities occurred, but correlations differed in different brain regions. Patterns of GM volume loss and Aβ aggregation in mild cognitive impairment and AD patients in frontal regions (Wirth et al., 2018).

### 4.1 Myelination

Taubert et al. showed an age-related myelin loss in the somatosensory and motor cortices as well as in thalamus nuclei (Taubert et al., 2020). Here, we reported a significant difference in the right hippocampus, indicating less myelin content in AD patients than healthy controls. At a more lenient statistical threshold (p <.001 uncorrected), there was a bilateral difference between groups indicating less myelin content in bilateral hippocampi in AD. This asymmetry of myelin deficits at the strictest statistical threshold can be related to our small sample size (only 24 participants within the AD continuum). These results agree with the previous findings on myelin deficits that lead to motor dysfunction, impaired cognitive functions, psychiatric disorders, and neurodegenerative disease (Chen, 2021; Wang et al., 2018). To assess the asymmetry of GM myelin alterations in AD, future studies with larger sample sizes and specifically designed analyses could provide more insights.

We also could show positive correlations between myelination, as indexed by MTsat map, and GM volume in AD, suggesting concurrence of GM loss and demyelination in the right para hippocampal area. Myelination was also highly correlated with synaptic density in the left fusiform. Our findings supports previous works that show myelin deficits are shown to relate to neurological setbacks such as motor dysfunction and neural degeneration (Wang et al., 2018). More generally, little is known about the GM myelination in the brain (Timmler and Simons, 2019), the specific relationship for myelination in gray matter is less well-studied. This is the first study to investigate myelination in GM using quantitative MRI method in Alzheimer’s disease.

### 4.2 Iron level

Studies on brain aging using the MPM approach demonstrated consistent findings on widespread negative correlation between age and MT saturation indicative for myelin loss paralleled by positive correlations between age and R2* in basal ganglia interpreted as iron content increases (Callaghan et al., 2014; Draganski et al., 2011; Taubert et al., 2020). We could not confirm the correlation between myelin and iron in our AD group. Previous in vivo studies (Duijn, 2017; Zeineh et al., 2015) showed that although iron content in the brain cortex and hippocampus is not affected by normal aging, in case of AD, accumulation of iron is observed in plaques, activated iron-containing microglia, and, in the most severe cases of AD, in the mid-cortical layers along myelinated fibers. Duijn et al also showed a difference in iron and myelin distribution in frontal cortex between the healthy controls and AD patients that are visible after development of AD pathological hallmarks (Duijn, 2017).

We didn’t observe any significant group difference (at voxel level) at the corrected statistical threshold for R2* maps indicative of iron content. However, at a more lenient statistical threshold (p <.001 uncorrected), we observed an increase in the iron content bilaterally in temporal lobe, and olfactory bulb, left hippocampus, and right supra marginal gyrus.

Su et al. reported that the iron content in the brain increased in one year compared to the baseline in AD participants and that iron accumulation was correlated with the neuropsychiatric behavior of participants (Su et al., 2016). Here, we showed a negative correlation between iron and GM volume in the right para hippocampal and left fusiform. The voxel-level comparison in the left hippocampus shows a significant mean difference between the groups.

### 4.3 GM volume

Voxel-based morphometry analysis showed bilateral difference between our groups, indicating loss in GM volume in AD patients consistently with the ROI comparisons, which are in accordance with aging studies that illustrate bilateral GM loss with respect to aging covering amygdala and hippocampus (Callaghan et al., 2014; Taubert et al., 2020). AD patients also showed reduced GM volume in left insula, olfactory bulb, and cingulate gyrus.

Although a correlation between memory and GM volume in hippocampus was expected (Dubois et al., 2007), here, we could not detect a significant correlation in our AD group perhaps due to the small size of the group.

### 4.4 Synaptic density

Our findings on the synaptic density, which is indexed by the Vt maps, show that synaptic loss occurred bilaterally in the hippocampus, where we could detect changes in the myelination and GM loss. Other regions that faced decreased synaptic density are the right and left thalamus. These results agree with previously published findings in (Bastin et al., 2020).

## 4.5 Concurrence of all image-derived biomarkers

The most important findings are provided by the multivariate results indicating that there is a significant co-occurrence of demyelination, iron accumulation, and GM and synaptic loss, in the hippocampal area bilaterally, left fusiform and temporal regions of AD brains compared to healthy brains. A recent study on aging indicates a complex pattern of atrophy, demyelination, and iron content change in the somatosensory cortex and motor area (Taubert et al., 2020).

Assuming that the probability maps are independent, the canonical vector information that measures the contribution rate of each modality within the multivariate model shows that in the clusters derived from statistical parametric map from mGLM, MTsat maps contribute the most to the observed difference, while R2* maps contribute the least in all clusters (see Figure 3). Currently, it is not possible to summarize and evaluate whether the profile of canonical vectors within a particular cluster is uniform or diverse. Furthermore, the interpretation of the findings is limited due to the lack of directionality of effects based on F-tests.

Comparisons of the original mean voxel values from the Z-transformed maps within the regions that the mGLM model could detect as significantly different between the groups suggest that most of these maps are independent. As a result, the canonical vectors are reliable measures for demonstrating the contribution rate of different image-derived biomarkers in the brain.

### 4.6 Limitations and conclusion

One limitation of the current study is the fact that only eight HC participants had a [18F]Flutemetamol PET scan to confirm Aβ-negativity. Although participants who did not undergo an amyloid PET were considered controls if they showed no or only minimal hippocampal atrophy on MRI, as assessed by visual inspection by a neurologist (Dubois et al., 2007), this does not preclude the possibility of the Aβ-positivity of some of the healthy controls. Despite this potential uncertainty, it is noteworthy that the AD group still showed a decrease in synaptic density in the hippocampus at voxel-level analysis. Another limitation is the lack of verification of the microstructural changes of the brain in different stages within the AD group, which consisted of subjects at various stages. There is still the need to conduct a longitudinal study from a large dataset that contains participants in different stages of AD. This would provide valuable insights into the chronobiology of microstructural changes and enable early detection of neurodegenerative diseases. Additionally, voxel-based quantification (VBQ) technique could be employed to preserve the quantitative value of the original qMRI maps (Draganski et al., 2011). Furthermore, future research should explore the use of quantitative susceptibility mapping (QSM) to overcome the limitations of R2*, as it is independent of water content, echo time, and field strength (Li et al., 2021).

Although MTsat maps are suggested to be proportional to the myelin content in the brain, they might also carry information on other macromolecular compounds in the brain that might reduce the specificity of MTsat to myelin alone. Inhomogeneous magnetization transfer (ihMT) is an alternative variant for MT that can address this issue (Munsch et al., 2021; Varma et al., 2020)

In conclusion, in this study, we compared individuals with Alzheimer’s pathology (amyloid positive) and healthy controls (HC) with multiple univariate and multivariate general linear models (GLMs) for three semi-quantitative maps (MTsat, R2*, and Vt) as well as gray matter (GM) volume. The complex interaction between various AD risk factors, such as myelination, iron accumulation, and neural degeneration, necessitates the use of a multivariate analytic approach. This approach is preferred over conducting multiple univariate comparisons for each modality studied, as it reduces the risk of false positives associated with multiple testing and increases sensitivity.

Although our study had a small sample size, the findings suggest that combining different image-derived AD risk factors in a multivariate analysis allows for the identification of specific brain regions where multiple neuropathological processes associated with the early stages of AD coincide. In addition to considering iron content and myelin using quantitative MRI, the novelty of our study lies in the comprehensive assessment of combined changes in AD biomarkers across the entire brain. In summary, our study underscores the importance of investigating Alzheimer’s disease (AD) from various pathological perspectives, as it is believed to involve a cascade of processes. By characterizing AD using non-invasive imaging techniques, we enhance the opportunity to detect and study the disease in its early stages, when interventions are most likely to be effective.

## Data Availability

All data produced in the present study are available upon reasonable request to the authors

## Acknowledgments

Funding: This work was supported by the ULiege Research Concerted Action (SLEEPDEM, grant 17/2109) and Walloon Region in the framework of the PIT program PROTHER-WAL under grant agreement No. 7289. [18F]flutemetamol was provided by GE Healthcare. CB is a Senior Research Associate at the F.R.S.-FNRS and CP is a Research Director at the F.R.S.-FNRS.

## Disclosure

The authors have no conflict of interest to declare.

## Notes

### Competing Interest Statement

The authors have declared no competing interest.

### Author Declarations

All procedures performed in studies involving human participants were approved by the Ethics Committee of the Liege University Hospital (Belgium), reference number EudraCT 2014-000286-50, and were in accordance with the 1964 Helsinki declaration and its later amendments or comparable ethical standards.”

### Summary of Updates

New correlation analyses are added to improve the discussion of the results. VBM analysis are added to both univariate and multivariate analyses, to consider the the effect of GM volume alteration.

