## Supplementary material for "Multimodal imaging of microstructural cerebral changes and loss of synaptic density in Alzheimer’s disease": Suplemantary data

Table sup-1. Pearson’s correlations between different maps in AD group with age and gender. Significant correlations are highlighted by * p < .05, ** p < .01, *** p < .001. Abbreviations: CI, confidence interval; MTsat, magnetization transfer saturation; Vt, total volume distribution; R2*, effective transverse relaxation rate; GMvol, gray matter volume.

|  | **Pearson's r** | **p-value** | **Lower 95% CI** | **Upper 95% CI** | **Fisher's z** |
| --- | --- | --- | --- | --- | --- |
| **Left hippocampus** | | | | | |
| **MTsat – Age** | -0.431 | 0.035 | -0.711 | -0.034 | -0.461 |
| **MTsat - Gender** | -0.258 | 0.224 | -0.599 | 0.163 | -0.264 |
| **R2* - Age** | -0.207 | 0.332 | -0.563 | 0.215 | -0.210 |
| **R2*- Gender** | -0.085 | 0.693 | -0.472 | 0.330 | -0.085 |
| **GMvol - Age** | -0.173 | 0.418 | -0.539 | 0.247 | -0.175 |
| **GMvol - Gender** | -0.591 | 0.002 | -0.803 | -0.246 | -0.679 |
| **Vt -Age** | -0.253 | 0.233 | -0.596 | 0.167 | -0.259 |
| **Vt - Gender** | -0.253 | 0.232 | -0.596 | 0.167 | -0.259 |
| **Right hippocampus** |  |  |  |  |  |
| **MTsat – Age** | -0.399 | 0.053 | -0.691 | 0.005 | -0.422 |
| **MTsat - Gender** | -0.445 | 0.029 | -0.719 | -0.051 | -0.479 |
| **R2* - Age** | -0.166 | 0.439 | -0.534 | 0.255 | -0.167 |
| **R2*- Gender** | -0.127 | 0.553 | -0.505 | 0.291 | -0.128 |
| **GMvol - Age** | -0.132 | 0.540 | -0.508 | 0.287 | -0.132 |
| **GMvol - Gender** | -0.385 | 0.064 | -0.682 | 0.022 | -0.405 |
| **Vt -Age** | -0.298 | 0.157 | -0.626 | 0.120 | -0.307 |
| **Vt - Gender** | -0.136 | 0.526 | -0.511 | 0.283 | -0.137 |
| **Right para hippocampal** | | | | | |
| **MTsat – Age** | -0.283 | 0.180 | -0.616 | 0.136 | -0.291 |
| **MTsat - Gender** | -0.394 | 0.057 | -0.688 | 0.012 | -0.416 |
| **R2* - Age** | 0.139 | 0.517 | -0.280 | 0.514 | 0.140 |
| **R2*- Gender** | 0.376 | 0.070 | -0.032 | 0.677 | 0.396 |
| **GMvol - Age** | -0.416 | 0.043 | -0.702 | -0.015 | -0.443 |
| **GMvol - Gender** | -0.537 | 0.007 | -0.773 | -0.171 | -0.600 |
| **Vt -Age** | -0.139 | 0.517 | -0.514 | 0.280 | -0.140 |
| **Vt - Gender** | -0.122 | 0.570 | -0.501 | 0.296 | -0.123 |
| **Left fusiform** | | | | | |
| **MTsat – Age** | -0.352 | 0.091 | -0.662 | 0.060 | -0.368 |
| **MTsat - Gender** | -0.503 | 0.012 | -0.753 | -0.125 | -0.553 |
| **R2* - Age** | 0.032 | 0.883 | -0.377 | 0.430 | 0.032 |
| **R2*- Gender** | 0.406 | 0.049 | 0.004 | 0.696 | 0.431 |
| **GMvol - Age** | -0.207 | 0.333 | -0.563 | 0.215 | -0.210 |
| **GMvol - Gender** | -0.559 | 0.004 | -0.786 | -0.202 | -0.632 |
| **Vt -Age** | -0.139 | 0.516 | -0.514 | 0.280 | -0.140 |
| **Vt - Gender** | -0.428 | 0.037 | -0.709 | -0.029 | -0.457 |
| **Left temporal** | | | | | |
| **MTsat – Age** | -0.631 | < .001 | -0.824 | -0.305 | -0.742 |
| **MTsat - Gender** | -0.395 | 0.056 | -0.689 | 0.010 | -0.418 |
| **R2* - Age** | -0.070 | 0.744 | -0.461 | 0.343 | -0.070 |
| **R2*- Gender** | 0.189 | 0.376 | -0.232 | 0.551 | 0.192 |
| **GMvol - Age** | -0.343 | 0.101 | -0.656 | 0.070 | -0.357 |
| **GMvol - Gender** | -0.239 | 0.261 | -0.586 | 0.182 | -0.244 |
| **Vt -Age** | -0.448 | 0.028 | -0.721 | -0.055 | -0.482 |
| **Vt - Gender** | -0.125 | 0.562 | -0.503 | 0.294 | -0.125 |

Tablesup- 2. Pearson’s correlations between different maps in HC group with age and gender. Significant correlations are highlighted by * p < .05, ** p < .01, *** p < .001. Abbreviations: CI, confidence interval; MTsat, magnetization transfer saturation; Vt, total volume distribution; R2*, effective transverse relaxation rate; GMvol, gray matter volume.

|  | **Pearson's r** | **p-value** | **Lower 95% CI** | **Upper 95% CI** | **Fisher's z** |
| --- | --- | --- | --- | --- | --- |
| **Left hippocampus** | | | | | |
| **MTsat – Age** | -0.663 | 0.002 | -0.859 | -0.299 | -0.798 |
| **MTsat - Gender** | 0.014 | 0.954 | -0.443 | 0.466 | 0.014 |
| **R2* - Age** | -0.266 | 0.271 | -0.643 | 0.214 | -0.273 |
| **R2*- Gender** | -0.224 | 0.356 | -0.616 | 0.256 | -0.228 |
| **GMvol - Age** | -0.378 | 0.111 | -0.710 | 0.092 | -0.397 |
| **GMvol - Gender** | -0.646 | 0.003 | -0.851 | -0.272 | -0.769 |
| **Vt -Age** | -0.114 | 0.642 | -0.540 | 0.359 | -0.115 |
| **Vt - Gender** | 0.101 | 0.680 | -0.370 | 0.531 | 0.102 |
| **Right hippocampus** | | | | | |
| **MTsat – Age** | -0.574 | 0.010 | -0.816 | -0.162 | -0.654 |
| **MTsat - Gender** | 0.019 | 0.938 | -0.439 | 0.469 | 0.019 |
| **R2* - Age** | -0.243 | 0.317 | -0.628 | 0.238 | -0.248 |
| **R2*- Gender** | -0.547 | 0.015 | -0.802 | -0.124 | -0.614 |
| **GMvol - Age** | -0.709 | < .001 | -0.880 | -0.376 | -0.885 |
| **GMvol - Gender** | -0.401 | 0.088 | -0.724 | 0.065 | -0.425 |
| **Vt -Age** | -0.163 | 0.504 | -0.575 | 0.314 | -0.165 |
| **Vt - Gender** | 0.055 | 0.822 | -0.409 | 0.497 | 0.055 |
| **Right para hippocampal** | | | | | |
| **MTsat – Age** | -0.176 | 0.470 | -0.584 | 0.302 | -0.178 |
| **MTsat - Gender** | -0.168 | 0.493 | -0.578 | 0.310 | -0.169 |
| **R2* - Age** | -0.207 | 0.396 | -0.604 | 0.273 | -0.210 |
| **R2*- Gender** | -0.572 | 0.011 | -0.814 | -0.159 | -0.650 |
| **GMvol - Age** | -0.530 | 0.020 | -0.793 | -0.100 | -0.590 |
| **GMvol - Gender** | -0.501 | 0.029 | -0.778 | -0.061 | -0.551 |
| **Vt -Age** | 0.013 | 0.959 | -0.444 | 0.464 | 0.013 |
| **Vt - Gender** | 0.075 | 0.760 | -0.393 | 0.512 | 0.075 |
| **Left fusiform** | | | | | |
| **MTsat – Age** | 0.094 | 0.702 | -0.376 | 0.526 | 0.094 |
| **MTsat - Gender** | -0.318 | 0.184 | -0.675 | 0.159 | -0.330 |
| **R2* - Age** | 0.117 | 0.633 | -0.356 | 0.542 | 0.118 |
| **R2*- Gender** | -0.540 | 0.017 | -0.798 | -0.114 | -0.604 |
| **GMvol - Age** | -0.668 | 0.002 | -0.861 | -0.306 | -0.806 |
| **GMvol - Gender** | -0.377 | 0.112 | -0.710 | 0.094 | -0.396 |
| **Vt -Age** | -0.267 | 0.269 | -0.643 | 0.213 | -0.274 |
| **Vt - Gender** | 0.021 | 0.933 | -0.438 | 0.470 | 0.021 |
| **Left temporal** | | | | | |
| **MTsat – Age** | -0.352 | 0.091 | -0.662 | 0.060 | -0.368 |
| **MTsat - Gender** | -0.503 | 0.012 | -0.753 | -0.125 | -0.553 |
| **R2* - Age** | 0.032 | 0.883 | -0.377 | 0.430 | 0.032 |
| **R2*- Gender** | 0.406 | 0.049 | 0.004 | 0.696 | 0.431 |
| **GMvol - Age** | -0.207 | 0.333 | -0.563 | 0.215 | -0.210 |
| **GMvol - Gender** | -0.559 | 0.004 | -0.786 | -0.202 | -0.632 |
| **Vt -Age** | -0.139 | 0.516 | -0.514 | 0.280 | -0.140 |
| **Vt - Gender** | -0.428 | 0.037 | -0.709 | -0.029 | -0.457 |

Table sup-3. Pearson’s correlations between different maps in HC group. Significant correlations are highlighted by * p < .05, ** p < .01, *** p < .001. Abbreviations: CI, confidence interval; MTsat, magnetization transfer saturation; Vt, total volume distribution; R2*, effective transverse relaxation rate, GMvol, gray matter volume.

|  | **Pearson's r** | **p-value** | **Lower 95% CI** | **Upper 95% CI** | **Fisher's z** | **VIF** |
| --- | --- | --- | --- | --- | --- | --- |
| **Left hippocampus** |  |  |  |  |  |  |
| **MTsat – R2*** | -0.031 | 0.899 | -0.479 | 0.429 | -0.031 | 0.999 |
| **MTsat - GMvol** | -0.082 | 0.738 | -0.517 | 0.386 | -0.082 | 0.993 |
| **MTsat - Vt** | 0.183 | 0.453 | -0.296 | 0.588 | 0.185 | 1.035 |
| **R2* - GMvol** | 0.538* | 0.018 | 0.110 | 0.797 | 0.601 | 1.407 |
| **R2*- Vt** | 0.253 | 0.297 | -0.228 | 0.634 | 0.258 | 1.068 |
| **GMvol - Vt** | -0.135 | 0.582 | -0.555 | 0.340 | -0.136 | 0.982 |
| **Right hippocampus** |  |  |  |  |  |  |
| **MTsat – R2*** | 0.144 | 0.556 | -0.332 | 0.562 | 0.145 | 0.979 |
| **MTsat - GMvol** | 0.284 | 0.238 | -0.195 | 0.654 | 0.292 | 0.919 |
| **MTsat - Vt** | 0.300 | 0.213 | -0.179 | 0.664 | 0.309 | 0.910 |
| **R2* - GMvol** | 0.193 | 0.427 | -0.286 | 0.595 | 0.196 | 0.963 |
| **R2*- Vt** | 0.250 | 0.302 | -0.231 | 0.632 | 0.255 | 0.937 |
| **GMvol - Vt** | -0.022 | 0.929 | -0.472 | 0.437 | -0.022 | 0.999 |
| **Right para hippocampal** |  |  |  |  |  |  |
| **MTsat – R2*** | 0.324 | 0.175 | -0.152 | 0.679 | 0.337 | 0.895 |
| **MTsat - GMvol** | 0.070 | 0.775 | -0.397 | 0.508 | 0.070 | 0.995 |
| **MTsat - Vt** | 0.170 | 0.486 | -0.308 | 0.580 | 0.172 | 0.971 |
| **R2* - GMvol** | 0.383 | 0.106 | -0.086 | 0.713 | 0.404 | 0.853 |
| **R2*- Vt** | 0.029 | 0.907 | -0.431 | 0.477 | 0.029 | 0.999 |
| **GMvol - Vt** | 0.128 | 0.601 | -0.346 | 0.550 | 0.129 | 0.984 |
| **Left fusiform** |  |  |  |  |  |  |
| **MTsat – R2*** | 0.471 | 0.042 | 0.022 | 0.762 | 0.512 | 0.778 |
| **MTsat - GMvol** | -0.180 | 0.462 | -0.586 | 0.299 | -0.182 | 0.968 |
| **MTsat - Vt** | -0.243 | 0.317 | -0.628 | 0.238 | -0.247 | 0.941 |
| **R2* - GMvol** | -0.190 | 0.436 | -0.593 | 0.289 | -0.192 | 0.964 |
| **R2*- Vt** | -0.215 | 0.376 | -0.610 | 0.265 | -0.219 | 0.954 |
| **GMvol - Vt** | 0.429 | 0.067 | -0.031 | 0.739 | 0.459 | 0.816 |
| **Left temporal** |  |  |  |  |  |  |
| **MTsat – R2*** | 0.361 | 0.129 | -0.111 | 0.700 | 0.378 | 0.870 |
| **MTsat - GMvol** | 0.144 | 0.557 | -0.332 | 0.561 | 0.145 | 0.979 |
| **MTsat - Vt** | -0.032 | 0.895 | -0.480 | 0.428 | -0.032 | 0.999 |
| **R2* - GMvol** | 0.752*** | < .001 | 0.453 | 0.899 | 0.978 | 0.434 |
| **R2*- Vt** | 0.307 | 0.201 | -0.171 | 0.668 | 0.317 | 0.906 |
| **GMvol - Vt** | 0.151 | 0.536 | -0.325 | 0.567 | 0.152 | 0.977 |
